# Gestational age-dependent decrease in fetal Hofbauer cells in placentas from pregnancies exposed to wildfire smoke in California

**DOI:** 10.1101/2023.01.11.23284125

**Authors:** Emilia Basilio, Nida Ozarslan, Sirirak Buarpung, Tarik Benmarhnia, Amy M. Padula, Joshua F. Robinson, Stephanie L. Gaw

**Author notes:** **Corresponding author:** Stephanie L. Gaw, MD PhD, Associate Professor, Division of Maternal-Fetal Medicine, Department of Obstetrics, Gynecology & Reproductive Sciences, University of California, San Francisco, 513 Parnassus Ave, Box 0556 16HSE, San Francisco, CA 94143. These authors jointly supervised this work (Amy M. Padula, Joshua F. Robinson, Stephanie L. Gaw). **Funding Sources:** This research was funded by the National Institute for Environmental Health Sciences (P30ES030284, R01ES031261). **Author Contribution:** Conceptualization, AMP, JFR, SLG; Writing – Original Draft Preparation, EB; Imaging, EB; Visualization, EB, NO, SB; Analysis, EB, Writing – Review & Editing, All authors. **Data Availability:** The data used to support the findings of this study are available from the corresponding author upon request. **Ethical Approval:** The University of California, San Francisco Committee on Human Research and Institutional Review Board approved this study (#21-33986 and #11-05530). **Consent:** All participants gave written informed consent. **Disclosure:** This work was presented in part during Oral Concurrent Session 3 at the Society for Maternal-Fetal Medicine 42^nd^Annual Meeting (Virtual, February 3, 2022).

## Abstract

**OBJECTIVE:** Wildfires are more common over the last decade and the frequency of wildfire events has been accelerated by climate change. The existing body of literature suggests that exposure to wildfire smoke during pregnancy contributes to adverse perinatal outcomes such as preterm birth and fetal growth restriction. We hypothesize that exposures to wildfire smoke and its constituents triggers a fetal inflammatory response which contributes to pathological changes that underlie these adverse pregnancy outcomes. In this study, we quantified the presence of fetal macrophages (i.e., Hofbauer cells) in human placentas obtained between 2018 and 2020 to assess the relationship between fetal immune status and wildfire exposure.

**STUDY DESIGN:** We collected placentas from pregnancies from two hospitals in San Francisco over a two-year period that included two severe major wildfires. The average particulate matter < 2.5 μm (PM_2.5_) or wildfire specific PM_2.5_ levels were estimated over the gestational duration of each sample. Immunostaining against CK7 and CD68 was performed to identify intravillous fetal Hofbauer cells. We assessed the gestational-age dependent relationship between placental CD68+ cell density and mean daily PM_2.5_ or wildfire-specific PM_2.5_ via linear regression and Welch’s t-test. Additionally, we compared placental CD68+ cell density with estimated peak wildfire exposures during the gestation to determine if timing of exposure during pregnancy may influence the occurrence of Hofbauer cells in the placenta.

**RESULTS:** The gestational ages ranged from 7-41 weeks (n = 67). The majority of samples were collected during one of two major wildfire events in Northern California (70%; n = 47). In general, we observed a significant inverse relationship between placental CD68 density and PM2.5 or wildfire specific PM2.5, however, these associations were only observed in first or second trimester samples, and not in term samples. For example, among first trimester samples (n=22), we observed lower mean CD68 density among samples likely to be exposed to wildfire events (*mean* = 1.42, *SD* = 0.8) as compared to those not exposed (*mean* = 3.73, *SD* = 1.983) (*p* = 0.0015). Based on our linear regression model results, we predicted that a one μg/m^3^ increase in daily mean wildfire PM_2.5_ was associated with a 0.457 decrease in CD68 density (ß =-0.457; 95% CI: -0.722, -0.193). This association was also significant for daily mean overall PM_2.5_, though smaller in magnitude (ß = -0.139; 95% CI: -0.218, -0.059).

**CONCLUSIONS:** Our results suggest that wildfire smoke exposures are associated with decreased presence of fetal Hofbauer cells in first and second trimester placentas, suggesting exposure may lead to impaired placental function via altered presence of fetal Hofbauer cells and changes in immune status.

## 1. Introduction

Wildfire events have intensified significantly, essentially quadrupling in the last four decades in the United States [1]. From 2018 to 2020 in California, over 6.4 million acres burned across the state [2]. Due to the burning of biomass and manmade structures, wildfire smoke contains a mixture of particles and chemicals including high levels of carbon monoxide, carbon dioxide, and particulate matter (PM) [3]. Wildfire smoke composition differs from typical air pollution because PM generated by wildfires is more toxic than non-fire period ambient PM secondary pollutants formed in atmospheric photochemistry reactions [3]. The contribution of wildfire smoke to PM_2.5_ (PM <2.5 microns in aerodynamic diameter) concentration in the US has grown substantially in the past 20 years. It accounts for nearly half of the overall PM_2.5_ exposure in western regions as compared to <20% a decade ago [4]. Approximately 70% of population in California experienced over 100 days of unhealthy air quality with PM_2.5_ levels in 2020 [5]. Wildfire smoke contains chemicals toxic to humans such as heavy metals (*e*.*g*., lead, cadmium, mercury) and polycyclic aromatic hydrocarbons (PAHs), especially those of low molecular weight (*e*.*g*., naphthalene, phenanthrene). These chemicals are found at greater amounts than in typical ambient air pollution sources [6]. In a recent review by our group, we found that emerging pregnancy studies (in human and animal) suggest that exposure to wildfire-specific air pollution may lead to adverse effects to the maternal-fetal unit via multiple pathways such as inflammation, oxidative stress, epigenetic changes in addition to metabolic, vascular, and endothelial dysregulation [7].

### 1.1 Inflammation from wildfire smoke exposure

Inflammation in pregnancy is strongly associated with adverse pregnancy outcomes which can have morbidity implications for the fetus and pregnant person [8,9]. Common pregnancy complications such as preterm birth, preeclampsia, and fetal growth restriction may result from altered balance of anti- and pro-inflammatory mediators [7]. Inflammatory responses are tightly regulated to promote a healthy pregnancy thus any alterations in inflammatory signals have been linked with early parturition resulting in preterm birth [10]. Higher levels of pro-inflammatory mediators in the maternal peripheral blood are associated with spontaneous preterm birth [11,12]. The activation of immune cells from environmental triggers can lead to a disruption of the homeostatic balance and could play a part in triggering preterm labor [13, 14].

While evidence supporting wildfire-specific inflammation during pregnancy is limited, growing evidence in animal and *in vitro* models suggests that wildfire smoke exposures modulate inflammatory pathways in maternal and fetal cells. In a first trimester trophoblast cell line, wood smoke exposures elevate secretion of IL-6, a major pro-inflammatory cytokine and led to decreased secretion of human chorionic gonadotropin and disrupted the membrane integrity of trophoblasts [15]. This study is supported by others showing that PM exposure from air pollution causes systemic inflammation [16] and the release of cytokines IL-6 and IL-8 in human bronchial epithelial cells and alveolar macrophages [17]. In addition, PAH exposures also hyperstimulate maternal immune cells in mouse models and exert a cytotoxic effect on human-derived macrophages [18,19].

The 2018 Camp fire was a major wildfire event in Northern California that destroyed nearly 19,000 structures and contained high levels of phthalates, lead, and zinc due to combustion of houses, cars, and other objects containing plastics [20]. During the 2018 Camp fire, rhesus monkeys that were exposed to wildfire smoke during the first trimester showed greater inflammation, blunted cortisol, and elevated C-reactive protein levels as compared to unexposed monkeys [21]. Rhesus macaques exposed to smoke during their infancy from the 2008 wildfire season in California were found to have differential expression of 172 genes in pathways critical for leukocyte extravasation signaling, CCR5 signaling in macrophages, and MIF regulation of innate immunity in nasal epithelium samples suggesting systemic inflammation [22]. While these initial studies suggest that devastating wildfire events may influence inflammation pathways, the effects of wildfire smoke on human pregnancy and the placenta are not yet understood.

### 1.2 The placental response to wildfire smoke exposure

The placenta serves several critical functions for embryonic and fetal growth. It acts as the primary metabolic barrier to environmental chemicals. Black carbon derived from air pollution is thought to transfer to maternal lung tissue, enter the systemic circulation, and translocate into placental tissue, accumulating in the trophoblast cell layer [23,24]. Many other toxic constituents of wildfire smoke are also known to accumulate in the placenta [25,26]. These findings suggest that the placental barrier is not impenetrable to PM and other environmental chemicals found in wildfire smoke, although it is unknown whether black carbon reaches the developing fetus directly or if this PM remains in the placenta [27]. These results are consistent with a recent *in vivo* study of human first trimester trophoblast cell line HTR-8 that was exposed to wood smoke [15]. PM was identified inside woodsmoke exposed cells and localized to the mitochondria and endoplasmic reticulum using transmission electron microscopy [15].

### 1.3 The response of fetal Hofbauer cells to wildfire smoke exposure

Monocytes/macrophages are principal cells involved in immune response [28]. Monocytes are leukocytes that originate from the hematopoietic stem cells of the bone marrow. They circulate in the blood with a lifespan of 1–2 days unless they are recruited to tissue in order to differentiate into macrophages [28]. Macrophages can originate from the differentiation of bone marrow-derived blood monocytes. The majority of these cells are derived from erythromyeloid progenitors arising from the embryonic yolk sac which survive until adulthood. Fetal Hofbauer cells (HBCs) are tissue macrophages localized within the stromal core of the placental villus and are the fetus’s primary immune cells at the maternal-fetal interface [29]. They are highly adaptive to the local microenvironment and are attributed to maintaining the balance between inflammation and tissue homeostasis, especially after infection or injury [29]. We hypothesize that black carbon and chemical contaminants of wildfire smoke modulate inflammatory signals in the human placenta leading to adverse pregnancy outcomes.

Therefore, in this study, we examined the presence of fetal Hofbauer cell populations, a critical cell population in fetal immune response, in human placentas of different gestational ages collected during major wildfire events in the San Francisco Bay Area from 2018 to 2020.

## 2. Materials and Methods

### 2.1 Participant Recruitment and Tissue Sampling

We collected placentas from uncomplicated singleton pregnancies exposed from November 2018 to November 2020 at the University of California San Francisco Betty Irene Women’s Hospital and Zuckerberg San Francisco General Hospital. Placental biopsies were obtained from the first trimester (GA <14 weeks; n=22), second trimester (GA 14-22 weeks; n=32), and term pregnancies (GA 37-42 weeks; n=13) during and after wildfire events (n=67). All placental samples from the first and second trimesters were collected shortly after participants underwent elective pregnancy terminations. Characteristics of these participants were not obtained since they were recruited anonymously. Biological samples were processed promptly after collection, within 1 hour of procedure or delivery. Full-thickness 1.0 cm^2^ placental biopsies were acquired within 1 hour after delivery, washed thoroughly in PBS and placed in 10% neutral buffered formalin (24-48 hours). After fixation steps, placental biopsies were transferred to 70% EtOH, embedded in paraffin and sectioned (4 *μ*m).

### 2.2 Estimation of wildfire smoke exposure

The average daily PM_2.5._ and wildfire-specific PM_2.5_ levels during the gestation were estimated from day of conception until day of delivery using the San Francisco ZIP Code 94110 [31]. Air levels of wildfire-specific PM_2.5_ were estimated using an established method described previously [32]. Briefly, we first determined daily concentrations of total PM from any source at the ZIP code level using ensemble models that integrated multiple machine learning algorithms (*e*.*g*., random forest, gradient boosting, and deep learning) and multiple predictor variables such as outdoor PM_2.5_ measurements from US EPA Air Quality System, aerosol optical depth, plume height, and meteorological variables (*e*.*g*., precipitation, temperature, wind speed and direction). We defined whether a pregnancy was “exposed” to a wildfire event using a cutoff of wildfire-specific PM_2.5_ > 5 *μ*g/m^3^ during any day of the gestation. We also assessed the average daily AQI, PM_2.5_ and wildfire PM_2.5_ experienced over the duration of the pregnancy as continuous variables. We additionally noted the number of days in each pregnancy that wildfire PM_2.5_ exceeded 5 μg/m^3^ to capture the intensity of a wildfire event.

### 2.3 Immunohistochemistry

Immunostaining against cytokeratin 7 (CK7) and pan-macrophage antigen CD68 was performed to identify trophoblasts and Hofbauer cells, respectively. First, samples were deparaffinized at room temperature. Antigen retrieval was performed with Uni-trieve at 70°C for 40 min. The blocking step was performed with 10% normal donkey serum in tris-buffered saline (TBS) with 1% bovine serum albumin BSA at room temperature for 2 hr. Next, the blocking solution was removed and samples were incubated with the primary antibodies rabbit anti-CD68 (ab213363; Abcam; 1:100) and rat anti-Cytokeratin7 (7D3; produced in the Fisher Lab [33], 1:50) and shielded from light overnight in a moist chamber at 4°C. The samples were then washed in TBS and 0.025% Triton X-100 and incubated with secondary antibodies, Alexa Flour 488 donkey anti-rabbit (ab150073; Abcam; 1:200) and Alexa Flour 594 donkey anti-rat (ab150156; Abcam; 1:200) for 1 hour at 37°C. Samples were then washed in TBS. The nuclei were stained using 40,60-diamidino-2-phenylindole (DAPI) in Vectashield solution (Vector Laboratories). The specimens were analyzed under a microscope and captured via Leica LAS X software (LEICA M80/DFC495). Ten representative villi per biopsy were examined in a blinded fashion by two members of the research team. Villous cross-sectional area was estimated using ImageJ software and cell counts were performed at 400X magnification. The total number of CD68+ cells were normalized against the total villous cross-sectional area to obtain a CD68+ cell density (**Figure 1**).

**Figure 1.**
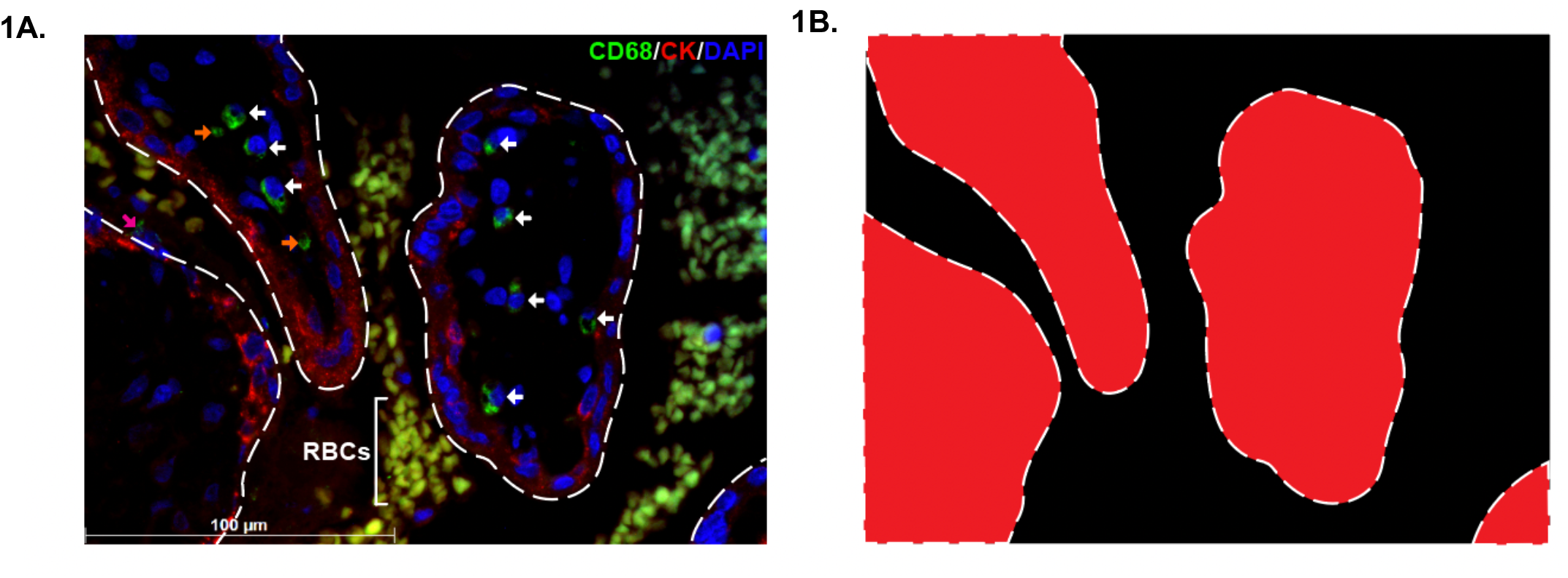
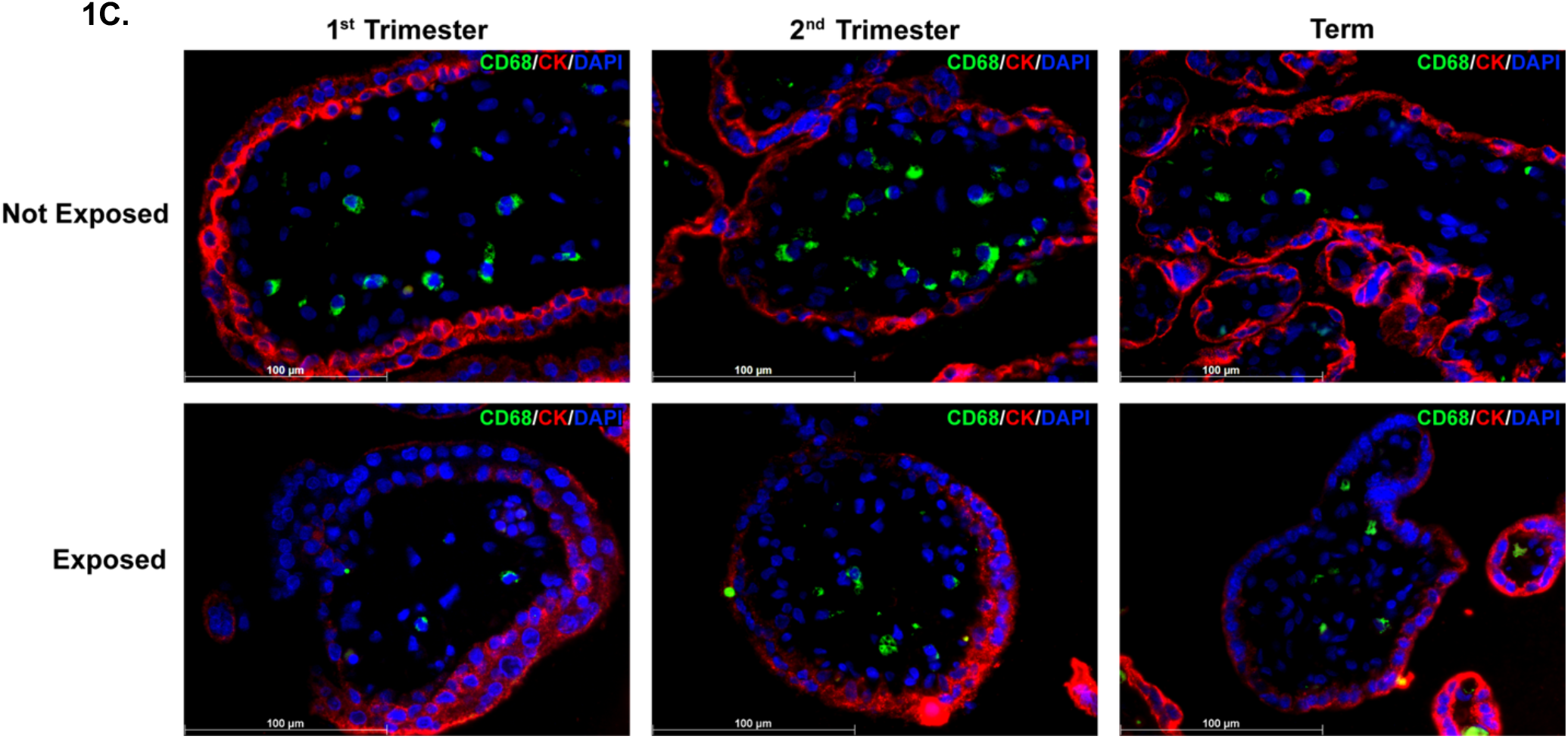
Quantification of Hofbauer cell density in placental villi. We used cell-specific antibodies, anti-CD68 (monocyte/macrophage) and anti-CK7 (trophoblast), to quantify the number of Hofbauer cells present in placental villi. The top panel represents the strategy used to identify and quantify density of Hofbauer cells (white arrow) in placental villi (400X magnification) (**1A** and **1B**). The orange arrow demonstrates an example of artifact which was not counted as a Hofbauer cell (no nuclei present); red-blood cells (RBCs) are noted by their autofluorescence and distinct appearance. Representative images of placental villi of each trimester that were exposed and unexposed to wildfires are shown above in (**1C**).

### 2.4 Statistical Analysis

We assessed the relationship of CD68 score with the daily average AQI, PM_2.5_, and wildfire specific PM_2.5_ via linear regression. Analyses were performed in Stata 16.1 and GraphPad Prism software (version 9.5.0). Two-tailed unpaired Welch’s t-test was performed to compare mean CD68 density against healthy controls among first trimester, second trimester, and term samples.

## 3. Results

The biopsies were examined with gestational ages ranging from 7-41 weeks (n = 67) (**Figure 2)**. The average daily PM_2.5_ in San Francisco varied during the study period with two distinct peaks during two major wildfire events the 2018 Camp Fire and the 2020 Glass Fire (**Figure 3)**.

**Figure 2.**
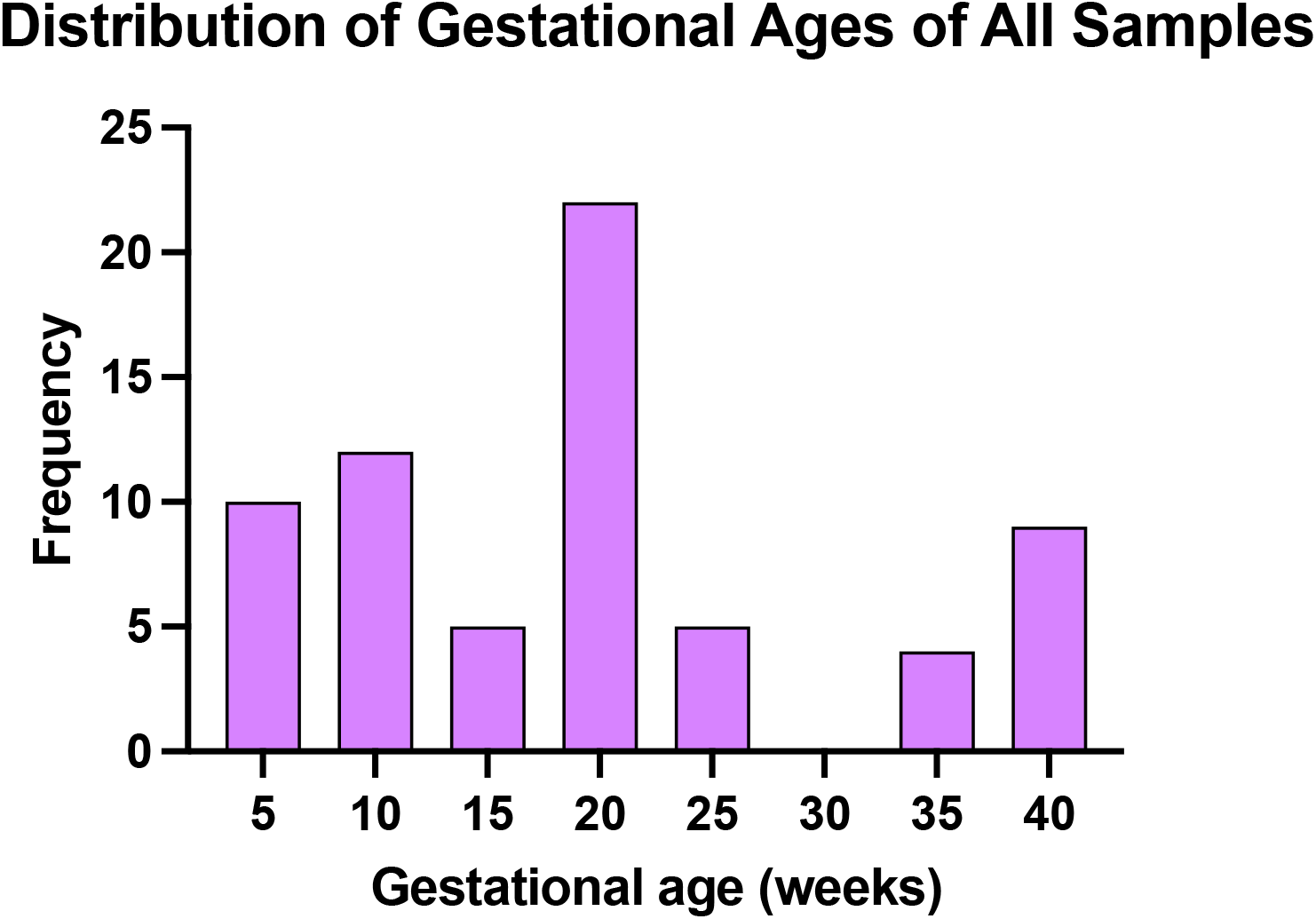
Frequency of samples at a given gestational age in weeks.

**Figure 3.**
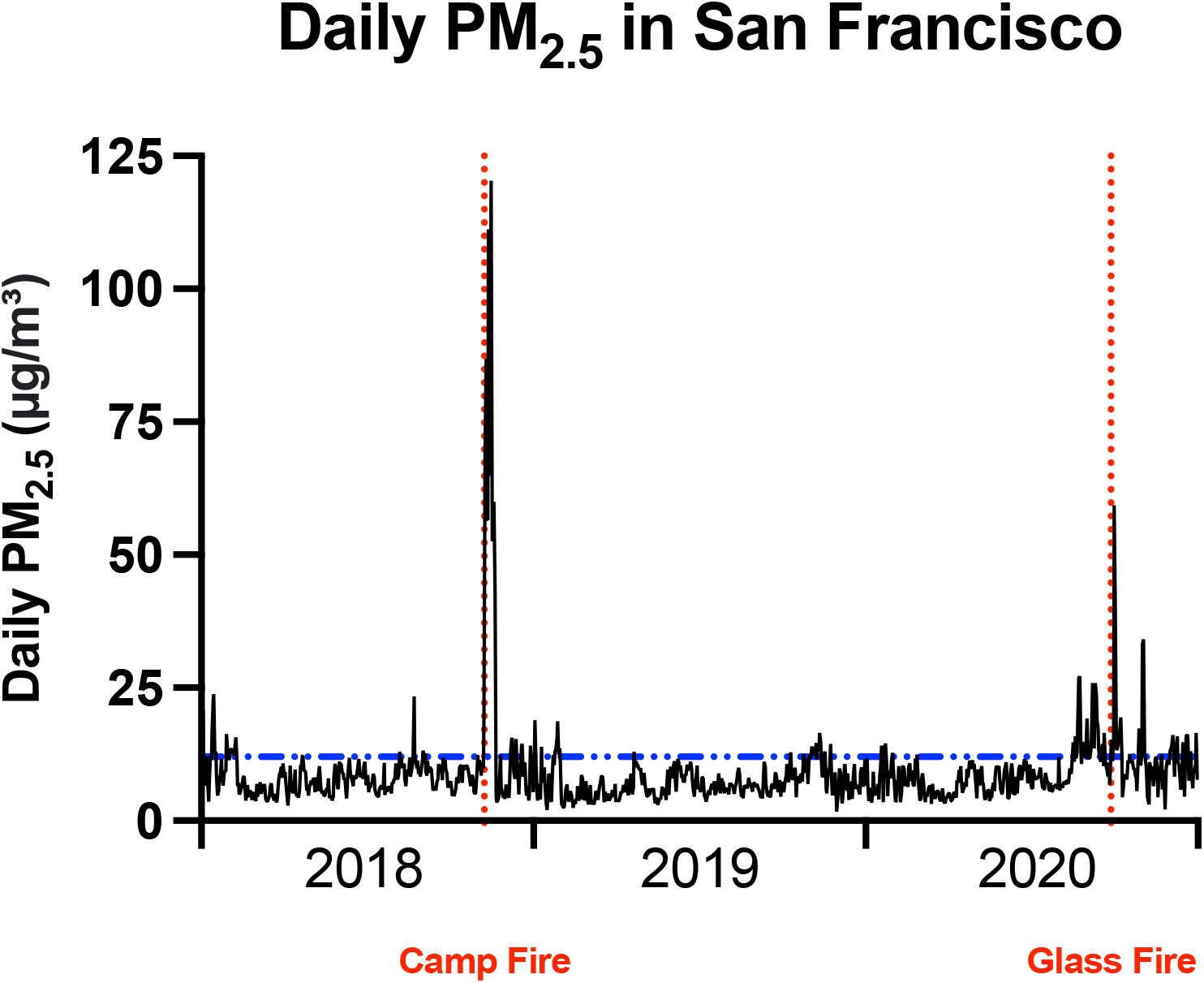
Daily PM_2.5_ estimates in San Francisco during 2018-2020. Placental samples were collected from pregnancies in San Francisco which overlapped with one of two major wildfires in California, the 2018 Camp Fire and the 2020 Glass Fire, red dotted lines. Blue dashed line denotes the threshold of ambient PM_2.5_ levels under which is healthy for all people (< 12 μg/m^3^).

The timing of the wildfire events occurred during different stages of the pregnancy as shown by the daily PM_2.5_ from San Francisco Zip Code 94110 plotted for each sample throughout the duration of each gestation for 2018 and 2020 (**Figure 4**). During the 2018 wildfire season, the first and second trimester samples were exposed to the Camp Fire. Term pregnancies in our study were exposed to the Glass Fire during the 2020 wildfire season. Both wildfire events resulted in multiple days’ worth of levels of PM_2.5_ that are considered unhealthy for sensitive populations like pregnant people.

**Figure 4.**
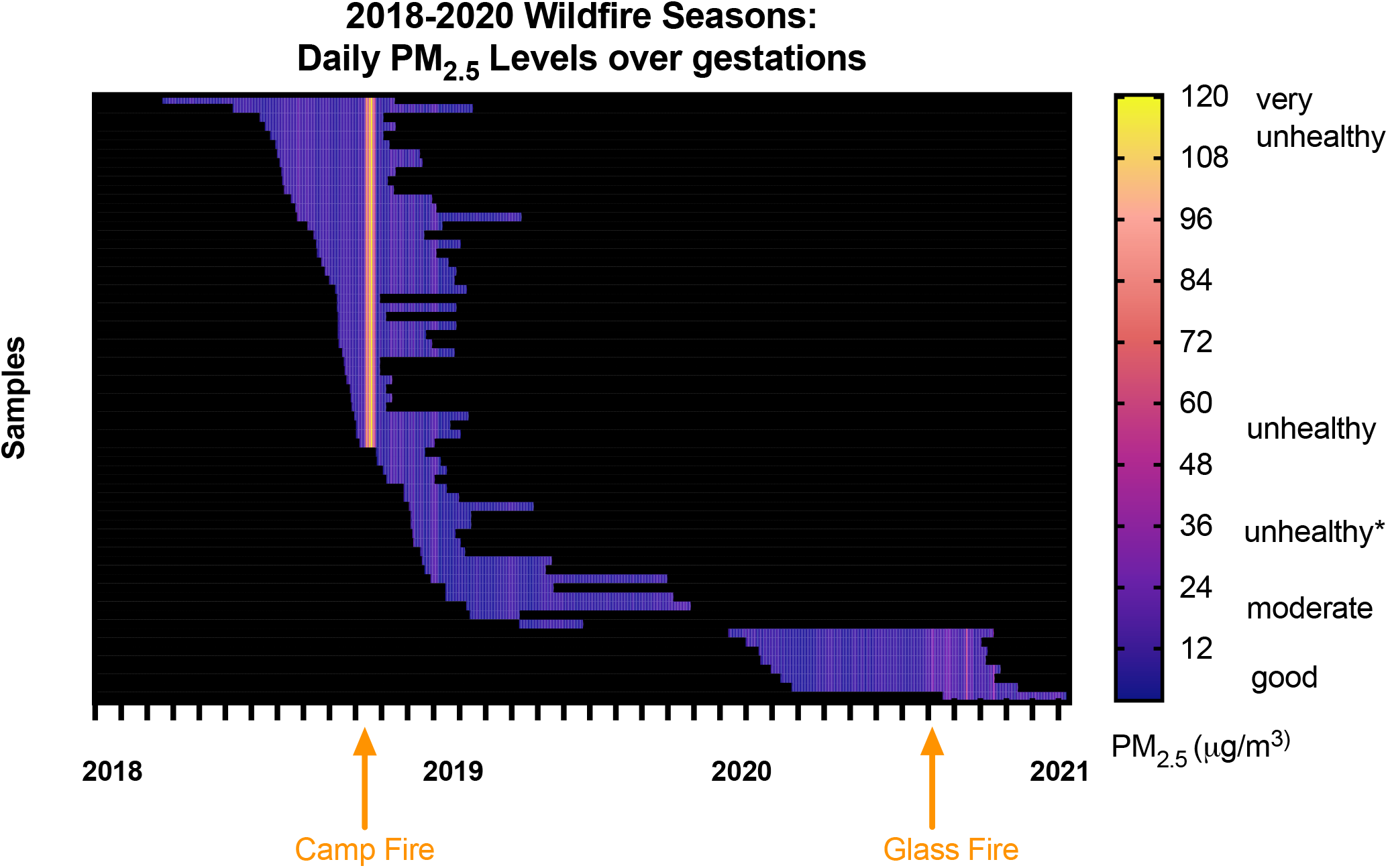
Daily PM_2.5_ values over each gestation during wildfire seasons in 2018 and 2020 are depicted above. We demonstrate the 2018 wildfire season that the first and second trimester samples were exposed to during which the Camp Fire occurred (orange arrow). We show the 2020 wildfire season that the term pregnancies were exposed to during which the Glass Fire occurred (orange arrow). Note that each wildfire event resulted in multiple days’ worth of levels of PM_2.5_ that are considered unhealthy for sensitive populations*, such as for pregnant persons (>35.4 μg/m^3^). **PM**_**2.5**_ **(μg/m**^**3**^**) Categorization:** good = <12; moderate 12.1-35.4; unhealthy for sensitive groups 35.5-55.4; unhealthy 55.5-150.4; very unhealthy 150.5-250.4; and hazardous >250.4.

The peak PM_2.5_ during the 2018 and 2020 wildfire seasons were 120.35 and 59.36 μg/m^3^, respectively. As mentioned previously, we defined “wildfire exposed” if a pregnancy experienced a peak wildfire specific PM_2.5_ >5 μg/m^3^ at any point during the pregnancy. In our sample (n = 67), the majority of pregnancies were wildfire exposed (70%, n = 47). In 2018, wildfire exposed first and second trimester pregnancies all experienced 11 consecutive days of wildfire specific PM_2.5_ > 5 μg/m^3^.

In 2020, the term wildfire exposed pregnancies experienced 2 consecutive days of wildfire specific PM_2.5_ above threshold. In the wildfire exposed group, on average each pregnant person experienced 30 days of moderate or worse quality defined by PM_2.5_ > 12 μg/m^3^ compared to 9 days for non-exposed persons.

The mean CD68 density among all wildfire exposed samples were not different than unexposed samples *p* = 0.738). There was no association between gestational age and mean CD68 density (R^2^ = 0.00254, *p* = 0.686) among all samples (**Figure 5A**). When stratified by wildfire exposure, there was a negative association between CD68 density and gestational age in unexposed pregnancies (R^2^ = 0.2755, *p* = 0.02), but not in exposed pregnancies (R^2^ = 0.00907, *p* = 0.524) (**Figure 5B**).

**Figure 5.**
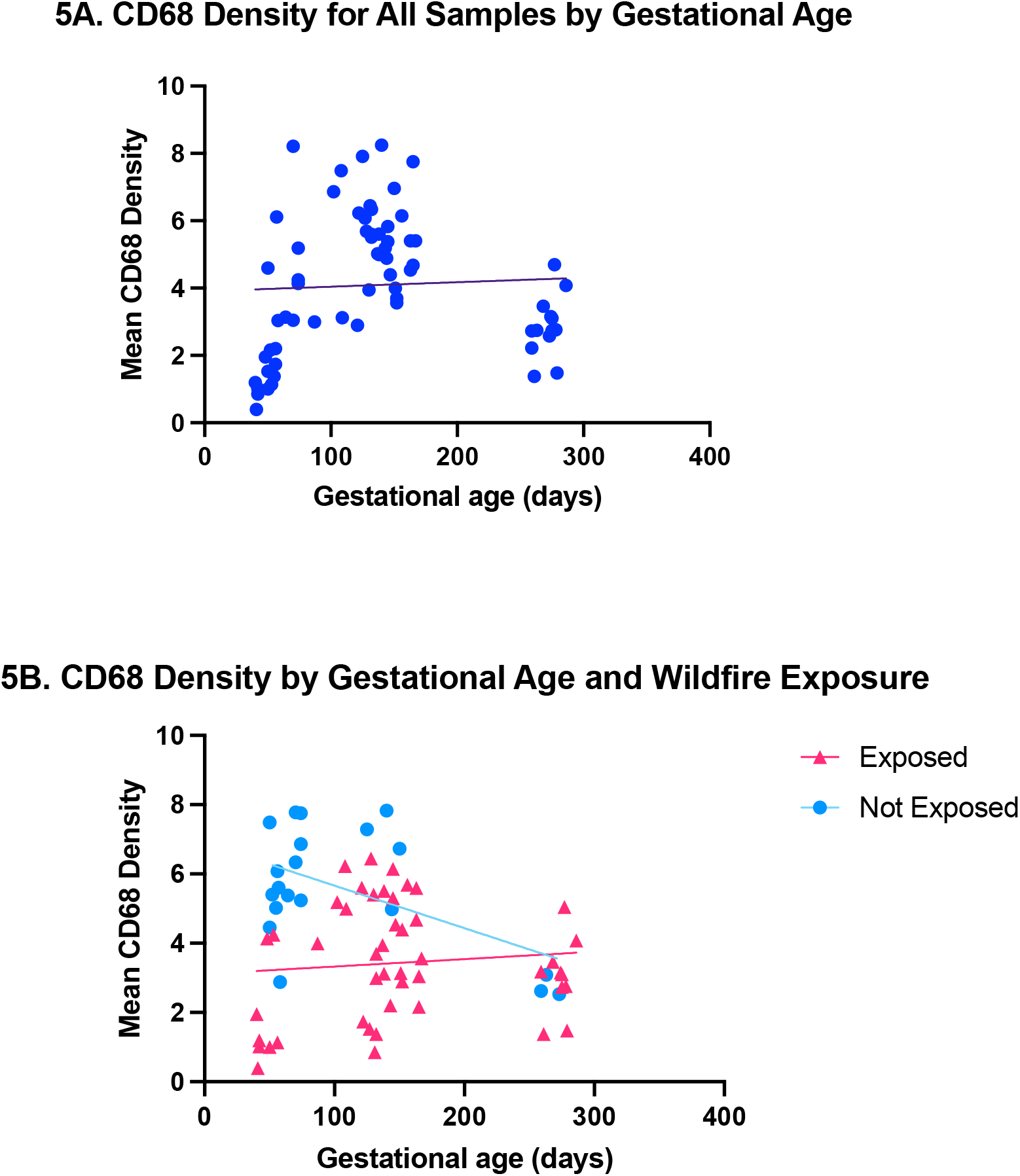
Density of intravillous Hofbauer cells in relation to gestational age and exposure to wildfire event. The density of Hofbauer cells in placental villi among all samples was not associated with gestational age (**5A**). This data suggests that exposed samples had a lower mean CD68 density throughout various gestational ages (**5B**).

We next performed linear regression performed to assess how wildfire specific PM_2.5_ and daily mean PM_2.5_ may impact CD68 density by trimester. We found that among first trimester samples, a one μg/m^3^ increase in daily mean wildfire PM_2.5_ was associated with a decrease in CD68 score (ß =-0.457; 95% CI: -0.722, -0.193). The magnitude of the association was smaller for daily mean overall PM_2.5_, though still statistically significant (ß = -0.139; 95% CI: -0.218, - 0.059). Among second trimester samples, a one μg/m3 increase in daily mean wildfire PM_2.5_ was associated with a decrease in CD68 score (ß = -0.680; 95% CI: -1.355, -0.004). The magnitude of the association was smaller for daily mean overall PM_2.5_ (ß= -0.167; 95% CI: -0.340, 0.005). For the term samples, there was no association between either wildfire specific PM_2.5_ or PM_2.5_ with the CD68 density (ß = 0.826, 95% CI: -0.677, 2.328 and ß = 0.106, 95% CI: -0.305, 0.516, respectively).

Across all samples at all gestational ages from 2018-2020, there was no difference in the mean CD68 density between wildfire exposed and unexposed samples (**Figure 6A)**. We next performed stratified analysis by trimester of collection to understand gestational-age dependent effects. Among first trimester samples (n = 22), mean CD68 density was lower among samples exposed to wildfire events (*mean* = 1.42, *SD* = 0.8) compared to those not exposed (*mean* = 3.73, *SD* = 1.983). The difference in mean CD68 density among wildfire exposed first trimester samples was significantly lower than unexposed samples, with the difference between means of - 2.313 (95% CI: -3.606, -1.022; *p* = 0.0015) (**Figure 6B**).

**Figure 6.**
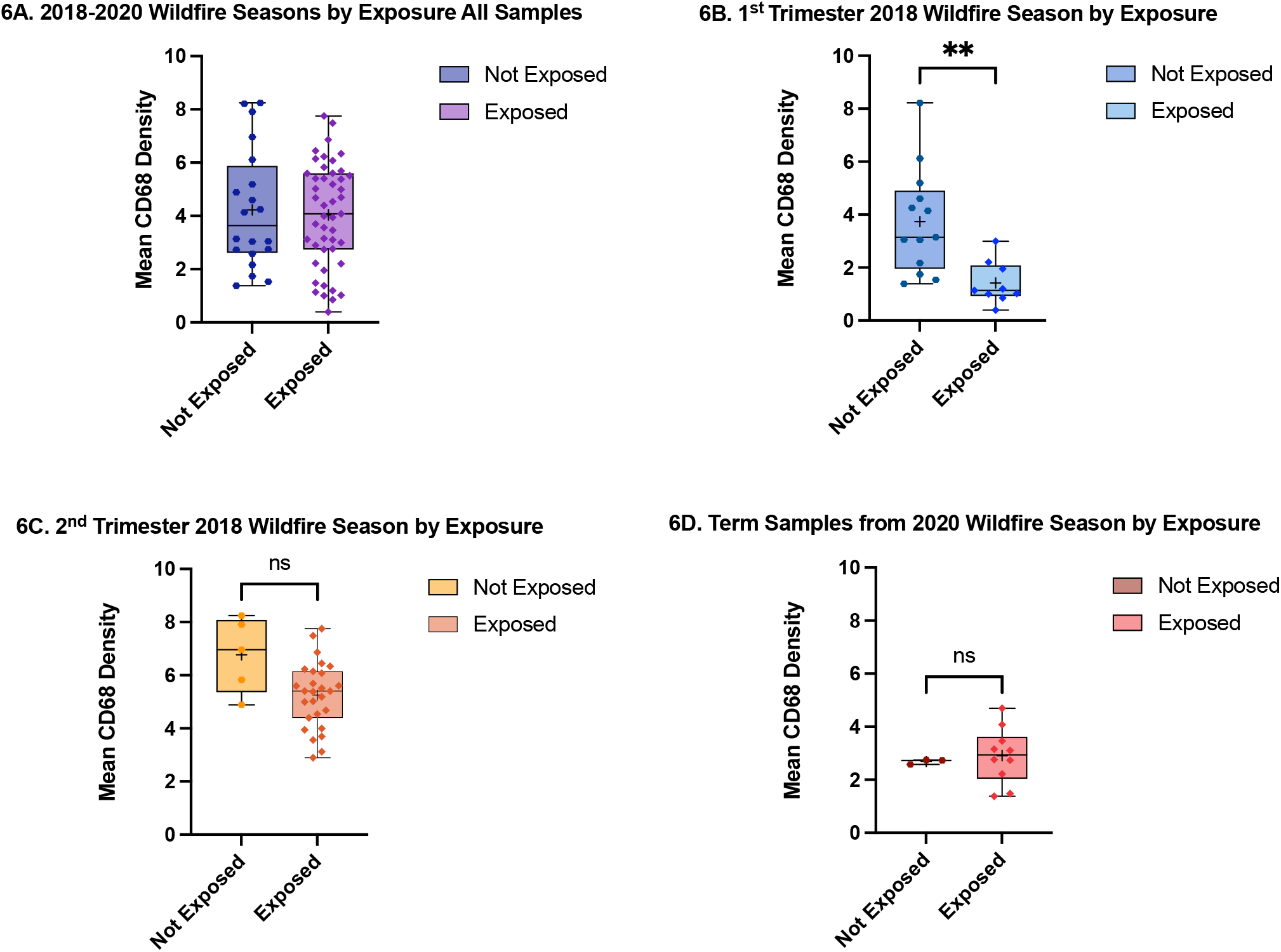
The mean CD68 density for all samples, first trimester, second trimester, and term pregnancies exposed and unexposed to wildfires from 2018-2020. Across all samples at all gestational ages from 2018-2020, there was no difference in the mean CD68 density between wildfire exposed and unexposed samples **(6A**). Among first trimester samples exposed to the 2018 Camp Fire, we note a statistically significant decrease in the mean CD68 density for exposed compared to unexposed samples (\*\**p* = 0.0015) **(6B)**. For second trimester samples exposed to the 2018 Camp fire, we note a decrease in the mean CD68 density for exposed compared to unexposed samples that is not statistically significant (*p* = 0.0729) (**6C**). Among term samples exposed to the 2020 Glass fire, there was no difference in mean CD68 density among exposed and unexposed samples (*p* = 0.520) (**6D**).

The difference in mean CD68 density among wildfire exposed first trimester samples was significantly lower than unexposed samples, with the difference between means of -2.313 (95% CI: -3.606, -1.022; *p* = 0.0015) (**Figure 6B**). Among second trimester (n = 32) and term samples (n=13), there were no statistically significant differences in the mean CD68 density with wildfire exposure (p=0.0729 and p=0.520, respectively; **Figure 6C and 6D**).

## 4. Discussion and Conclusions

This is the first study, to our knowledge, to characterize the density of fetal HBCs in placentas acquired during major wildfire events. Our data show that wildfire smoke exposure during pregnancy is associated with decreased quantity of placental HBCs during the first trimester and suggests a similar trend in the second trimester. Our data supports the hypothesis that wildfire smoke PM influences human placental immune response.

These findings provide the first evidence that wildfire smoke exposure in pregnancy may dysregulate placental immune status, particularly with exposures earlier in pregnancy. These results are concerning and build on previous examples that have shown chemical components of wildfire smoke such as polycyclic aromatic hydrocarbons (PAHs) to act as contribute to an immunosuppressive effect on macrophages [19]. There is little data on the composition of wildfires from distinct wildfire events. While our results are in contrast to other studies that have suggested a pro-inflammatory effect of wildfire smoke exposure in non-human primates, this may reflect species-specific responses to exposure [21]. The primate placenta is structurally distinct from the human placenta. One possible explanation are uncharacterized differences in the chemical composition of wildfire smoke that could be unique to the duration, location, and weather conditions during a wildfire event. There are several limitations to this study, including that we used outdoor air quality readings to define daily exposure. We did not have individual level exposure data which would help inform how participants in this study may have been differentially exposed to wildfire smoke or data regarding behavioral changes participants may have done to mitigate their exposures, such as using a high-quality mask while outdoors or use of in-home air purifiers. Additionally, we did not have clinical outcomes available for each participant to further characterize our sample population. The strengths of this study include that this is the first study to obtain placental samples over multiple time points in pregnancies that were exposed to significant recent wildfire events in California.

Future investigations, both *in vitro* and *in vivo* will be critical to closely examine if wildfire smoke exposures have other negative impacts on other cell lineages as well. Understanding how placental function is altered and the mechanisms by which wildfire smoke exposure may cause dysfunction will guide future therapeutics and may one day help mitigate adverse pregnancy outcomes that arise from chemical toxicants found in wildfire smoke.

## Data Availability

The data used to support the findings of this study are available from the corresponding author upon request.

## Abbreviations

GA: Gestational age
PM: Particulate matter
PM_2.5_: Particulate Matter <2.5 μm

## Acknowledgements

The authors would like to acknowledge all the participants who donated samples to this research. We would like to acknowledge Dr. David A. Ramos for assistance with image processing.

